# Effect of corticosteroid treatment on 1376 hospitalized COVID-19 patients. A cohort study.

**DOI:** 10.1101/2020.07.17.20155994

**Authors:** Filippo Albani, Federica Fusina, Enza Granato, Cristina Capotosto, Claudia Ceracchi, Riccardo Gargaruti, Giovanni Santangelo, Luca Schiavone, Maria Salvatrice Taranto, Cinzia Tosati, Elena Vavassori, Giuseppe Natalini

**Author notes:** **Corresponding Author** Federica Fusina, Department of Anesthesia, Intensive Care and Pain medicine, via Bissolati, 57, Brescia, 25124, IT.

## Abstract

**Background:** Since the start of the novel coronavirus 2019 (COVID-19) pandemic, corticosteroid use has been the subject of debate. The available evidence is uncertain, and knowledge on the subject is evolving. The aim of our cohort study was to evaluate the association between corticosteroid therapy and hospital mortality, in patients hospitalized with COVID-19 after balancing for possible confounders.

**Results:** One thousand four hundred forty four patients were admitted to our hospital with a positive RT-PCR test for SARS-CoV-2, 559 patients (39%) were exposed to corticosteroids during hospital stay, 844 (61%) were not exposed to corticosteroids.In the cohort of patients exposed to corticosteroids, 171 (30.6%) died. In the cohort of patients not exposed to corticosteroids, 183 (21.7%) died (unadjusted p <0.001). Nonetheless, exposure to corticosteroids was not associated with in-hospital mortality after balancing with overlap weight propensity score (adjusted p = 0.25). Patients in the corticosteroids cohort had reduced risk of ICU admission (adjusted p <0.001).

**Conclusions:** Treatment with corticosteroids did not affect hospital mortality in patients with COVID-19 after balancing for confounders. A possible advantage of corticosteroid therapy was to reduce Intensive Care Unit admission, which could be useful in reducing pressure on the Intensive Care Units in times of limited resources, as during the COVID-19 pandemic.

## Introduction

In December 2019 a cluster of atypical pneumonia was observed in Wuhan, China. A new coronavirus, later called Severe Acute Respiratory Syndrome Coronavirus 2 (SARS-CoV 2), was identified as responsible for the disease,^1^ which subsequently spread to the rest of the world. More than two thousand patients presented to our Emergency Department with a positive RT-PCR test for SARS-CoV 2 during the novel coronavirus disease (COVID-19) pandemic, from 20^th^ February to 10^th^ May. One thousand four hundred and forty three of them were admitted to the hospital.

Many pharmacological therapies have been attempted despite the absence of evidence supporting their efficacy, among them corticosteroids.

Initially the World Health organization discouraged the use of corticosteroids for the treatment of COVID-19 because their administration to patients with SARS had shown no survival benefit and possible harms (avascular necrosis, psychosis, diabetes, and delayed viral clearance). ^2–4^ The interest in corticosteroids was renewed by the observation that their use was associated with a mortality reduction in COVID-19 patients in China,^5^ and recommendations after this report included the use of steroids in patients with COVID-19 and ARDS as a weak recommendation.^6^

This uncertain and evolving evidence concerning the use of corticosteroids had, as a consequence, their inconstant use in COVID-19 patients. In our hospital they were used in about 40% of patients.

Gathering evidence on the effect of corticosteroids treatment in COVID-19 is crucial, because the infection rates of other human coronaviruses have a typical wave pattern with the pick in autumn\winter^7^ and a second surge of the disease could be possible.

The aim of our cohort study was to evaluate the association between corticosteroid therapy and hospital mortality in patients hospitalized with COVID-19 after balancing for possible confounders.

## Methods

The setting for this observational cohort study was a 600 bed tertiary care hospital located in Brescia (Northern Italy). Data was extracted from the electronic medical charts of consecutive patients who had been admitted during the pandemic crisis of Severe Acute Respiratory Syndrome CoronaVirus-2 (SARS-CoV-2), between February 20^th^ and May 10^th^ 2020. The study protocol was approved by the Ethical Committee of Brescia (NP-4210). All research was conducted in accordance with Declaration of Helsinki.

Clinical, demographic and laboratory data from all adult patients who were admitted to the hospital and had a positive test for SARS-CoV2 from biological material were recorded at admission. Tests were conducted with Real Time Polymerase-Chain-Reaction assay (RT-PCR).

Patients younger than 18 years or with no available outcome at the time of analysis were excluded from the study. Patients were followed up until discharge.

The patients were classified into two cohorts, based on exposure to corticosteroids during hospitalization.

In-hospital mortality was the primary outcome. The rate of admission to the Intensive Care Unit was the secondary outcome. Patients were admitted in ICU if they had, during Non Invasive Ventilation: PaO2/FiO2 < 100 mmHg, dyspnea, tachypnea (more than 40 breaths per minute), tidal volume greater than 10 ml/kg.

One thousand three hundred seventy seven patients were needed to obtain a power of 85% to detect an Odds Ratio 0.72, assuming mortality was 25% in those receiving corticosteroids, with a fraction of patients treated of 1:3 and alpha=0.05^8^. The 12 weeks follow up ensured 97% of patients were discharged or had died by the end of the study period. Continuous variables were expressed as mean (standard deviation) or as median (1^st^, 3^rd^ quartile) and factor variables as count (percentage). Bivariate analysis of outcome was realized on identified a priori variables with Fisher test for factorial variable and t-test and Mann-Whitney test for continuous ones.

A overlap weight propensity score for treatment allocation was estimated from a multivariable model containing patient age, sex, PaO_2_/FiO_2_, lactate, C Reactive Protein, Platelets, ICU admission and treatment with corticosteroids, enoxaparin, azithromycin or hydroxychloroquine. Dose of hydrocortisone and methylprednisolone were converted to equivalent doses of dexamethasone.^9^

The overlap weight propensity score was then applied to a logistic regression modelling the primary outcome (in-hospital mortality)^10^. Odds Ratio with 95% Confidence Interval (OR, 95%CI) are reported. The same was performed for OR of ICU admission, considering only corticosteroids treatment received on the wards. Missing values were assessed and replaced by mean substitution. If a statistical significant association was found, the E-values for the point estimate and the confidence interval limit closer to the null were computed, to assess the possible effect of unmeasured confounder.^11^ Three sensitivity analyses were planned: one taking into account the quartiles of timing of hospital admission, one excluding patients admitted to intensive care, another analyzing complete cases. Significance was evaluated at α = 0.05 and all testing was 2-sided. Statistical analysis was performed using R Studio software version 4.0.0 (R Core Team, 2014) and packages ‘psw’ and ‘E-value’ for overlap weight propensity score and calculation of E-value. Recorded data from this cohort has been used to evaluate effect of other pharmacological intervention in COVID 19, manuscripts at present are undergoing peer-review.

## Results

One thousand four hundred and forty three patients were admitted to our hospital with a positive RT-PCR test for SARS-CoV-2. Five hundred and fifty nine patients (39%) were exposed to corticosteroids during hospital stay, 844 (61%) were not exposed to corticosteroids. Twentyseven patients were excluded from the analysis (one patient was younger than 18 years, and 26 patients were still hospitalized at the time of the analysis). Median equivalent doses of dexamethasone dose (cumulative dose/days of therapy) was 10 mg (1st-3rd quartile 4.5-20). Patients who were never admitted to ICU received equivalent doses of dexamethasone of 8 mg (4-16.1). Duration of corticosteroid therapy was longer in survivors than in patients who died: 6 (4-10) days vs 4 (1-7), respectively (p <0.001).

Clinical and laboratory characteristics in the cohort of patients exposed and not exposed to corticosteroids are shown in Table 1. Age was not significantly different between the two cohorts, while Body Mass Index was higher in patients treated with corticosteroids. Arterial hypertension was more frequent in the corticosteroid group, while diabetes mellitus was not significantly different between the two groups. Patients in the corticosteroids cohort were more hypoxic and the inflammation markers (CRP and LDH) were higher, moreover lymphocytopenia was more severe. PaCO_2_ was not different in the two cohorts. One hundred seventy one patients died (30.6%) in the cohort of patients exposed to corticosteroids, and 183 (21.7%) died in the cohort of patients not exposed to corticosteroids (unadjusted p <0.001). Fifty six patients (11.5%) were admitted to ICU in the cohort of patients exposed to corticosteroids on the ward vs 131 (14.4%) who were not treated with corticosteroids in the ward (unadjusted p =0.15).

**Table 1.**

|  |  | not exposed to<br>corticosteroids | exposed to<br>corticosteroids | *p* value |
| --- | --- | --- | --- | --- |
| Patients | N (%) | 844 (60) | 559 (40) |  |
| Age | years | 68.5 (15.1) | 68.7 (11.5) | 0.853 |
| Body Mass Index | kg*m<sup>-2</sup> | 26.4 (5.0) | 27.3 (4.9) | 0.001 |
| Female | N (%) | 286 (33.9) | 193 (34.5) | 0.849 |
| Arterial hypertension | N (%) | 281 (33.3) | 218 (39.0) | 0.033 |
| Diabetes mellitus | N (%) | 164 (19.4) | 125 (22.4) | 0.207 |
| PaO<sub>2</sub> | mmHg | 59.9 (23.8) | 53.2 (18.5) | <0.001 |
| pH |  | 7.47 (0.06) | 7.48 (0.07) | 0.003 |
| PaCO<sub>2</sub> | mmHg | 35.2 (7.7) | 34.8 (8.5) | 0.356 |
| PaO<sub>2</sub>/FiO<sub>2</sub> | mmHg | 269.0 (75.1) | 235.2 (74.6) | <0.001 |
| HCO<sub>3</sub><sup>-</sup> | mmol*L<sup>-1</sup> | 24.7 (4.0) | 24.8 (3.7) | 0.551 |
| Lactate | mmol*L<sup>-1</sup> | 1.3 (1.0) | 1.4 (1.2) | 0.193 |
| Leukocytes | 10<sup>9</sup>*L<sup>-1</sup> | 8.0 (4.1) | 8.6 (4.4) | 0.009 |
| Lymphocytes | $10^9 \text{L}^{-1}$ | 1.2 (1.1) | 1.0 (1.0) | 0.028 |
| Platelets | $10^9 \text{L}^{-1}$ | 196.8 (91.7) | 203.3 (93.7) | 0.221 |
| C-reactive Protein | $\text{mg} \cdot \text{L}^{-1}$ | 98.6 (78.3) | 137.8 (93.1) | <0.001 |
| Lactate Dehydrogenase | $\text{U} \cdot \text{L}^{-1}$ | 442.6 (248.6) | 567.6 (224.6) | 0.030 |

Demographics, comorbidities, laboratory data and outcomes of the study subjects. Factor variables are expressed as count (%), continuous variables as mean (standard deviation) or as median (interquartile range). Fisher test was used for factorial variables and t-test and Mann-Whitney test for continuous ones. BMI, body mass index; $\text{PaO}_2$ partial pressure of arterial oxygen; $\text{PaCO}_2$ partial pressure of arterial carbon dioxide; $\text{PaO}_2/\text{FiO}_2$ , partial pressure of arterial oxygen to the fraction of inspired oxygen

All the recorded variables were associated with mortality in the bivariate analysis except for Body Mass Index, history of arterial hypertension or diabetes mellitus and PaCO_2_, as shown in Table 2.

**Table 2.**

|  |  | Survivors | Non-survivors | *p* value |
| --- | --- | --- | --- | --- |
| Patients | N (%) | 1022 (72.8) | 354 (25.2) |  |
| Age | years | 65.8 (14.0) | 76.4 (9.7) | <0.001 |
| Body Mass Index | kg*m<sup>-2</sup> | 26.6 (4.8) | 27.2 (5.3) | 0.071 |
| Female | N (%) | 374 (36.6) | 93 (26.3) | 0.001 |
| Arterial hypertension | N (%) | 377 (36.9) | 122 (34.5) | 0.451 |
| Diabetes mellitus | N (%) | 204 (20.0) | 85 (24.0) | 0.124 |
| PaO<sub>2</sub> | mmHg | 59.4 (19.8) | 50.9 (25.6) | <0.001 |
| PaCO<sub>2</sub> | mmHg | 35.0 (6.9) | 35.0 (10.2) | 0.914 |
| PaO<sub>2</sub>/FiO<sub>2</sub> | mmHg | 271.2 (69.5) | 210.3 (76.8) | <0.001 |
| pH |  | 7.48 (0.06) | 7.46 (0.07) | <0.001 |
| HCO<sub>3</sub><sup>-</sup> | mmol*L<sup>-1</sup> | 25.1 (3.5) | 23.9 (4.5) | <0.001 |
| Lactate | mmol*L<sup>-1</sup> | 1.2 (0.7) | 1.8 (1.5) | <0.001 |
| Leukocytes | 10<sup>9</sup>*L<sup>-1</sup> | 8.0 (3.7) | 8.8 (5.3) | 0.002 |
| Lymphocytes | $10^9 \cdot L^{-1}$ | 1.2 (1.0) | 0.9 (1.1) | <0.001 |
| Platelets | $10^9 \cdot L^{-1}$ | 205.5 (91.7) | 181.2 (87.7) | <0.001 |
| C-reactive Protein | $mg \cdot L^{-1}$ | 100.8 (81.8) | 154.6 (89.2) | <0.001 |
| Lactate Dehydrogenase | $U \cdot L^{-1}$ | 479.7 (218.4) | 658.2 (279.8) | 0.011 |

Demographics, comorbidities, laboratory data and outcomes of survivors and nonsurvivors. Factor variables are expressed as count (%), continuous variables as mean (standard deviation) or as median (interquartile range). Fisher test was used for factorial variables and t-test and Mann-Whitney test for continuous ones. BMI, body mass index; PaO<sub>2</sub> partial pressure of arterial oxygen; PaCO<sub>2</sub> partial pressure of arterial carbon dioxide; PaO<sub>2</sub>/FiO<sub>2</sub>, partial pressure of arterial oxygen to the fraction of inspired oxygen.

Covariate balancing with overlap weight propensity score is shown in Figure 1 expressed as standardized mean difference which became zero after balancing.

**Figure 1.**
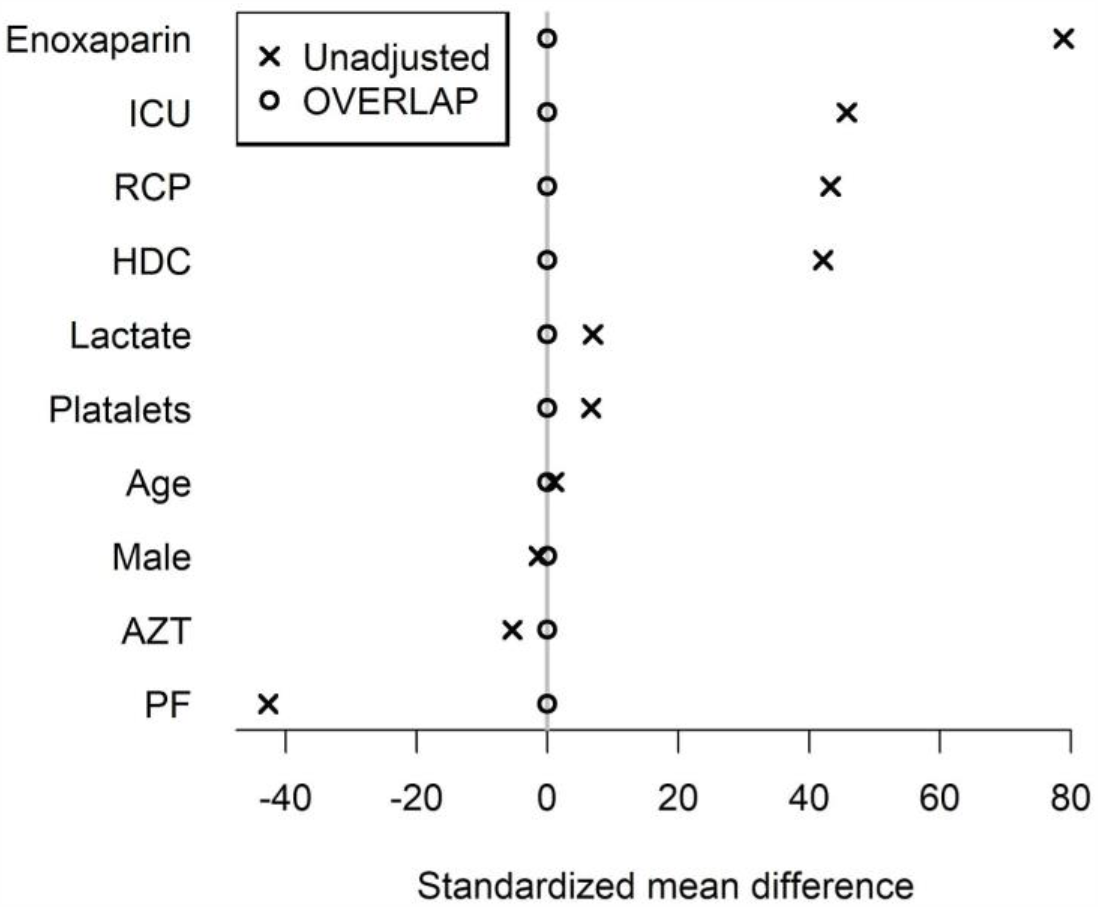
Covariance balance plot before and after overlap weights propensity matching for patients treated with corticosteroids and patients not treated with corticosteroid, expressed with standardized mean difference for comparability. ICU, Intensive Care Unit admission; RCP, Reactive C Protein; HDC, treatment with Hydroxychloroquine; AZT, treatment with Azithromycin; PF, PaO_2_/FiO_2_ ratio.

Table 3 shows that the exposure to corticosteroids was not associated with in-hospital mortality after balancing with overlap weight propensity score. Patients in the corticosteroids cohort had reduced risk of ICU admission and E-value of 2.26 (1.76 upper limit of C.I.). Planned sensitivity analyses were in agreement with these findings.

**Table 3.**
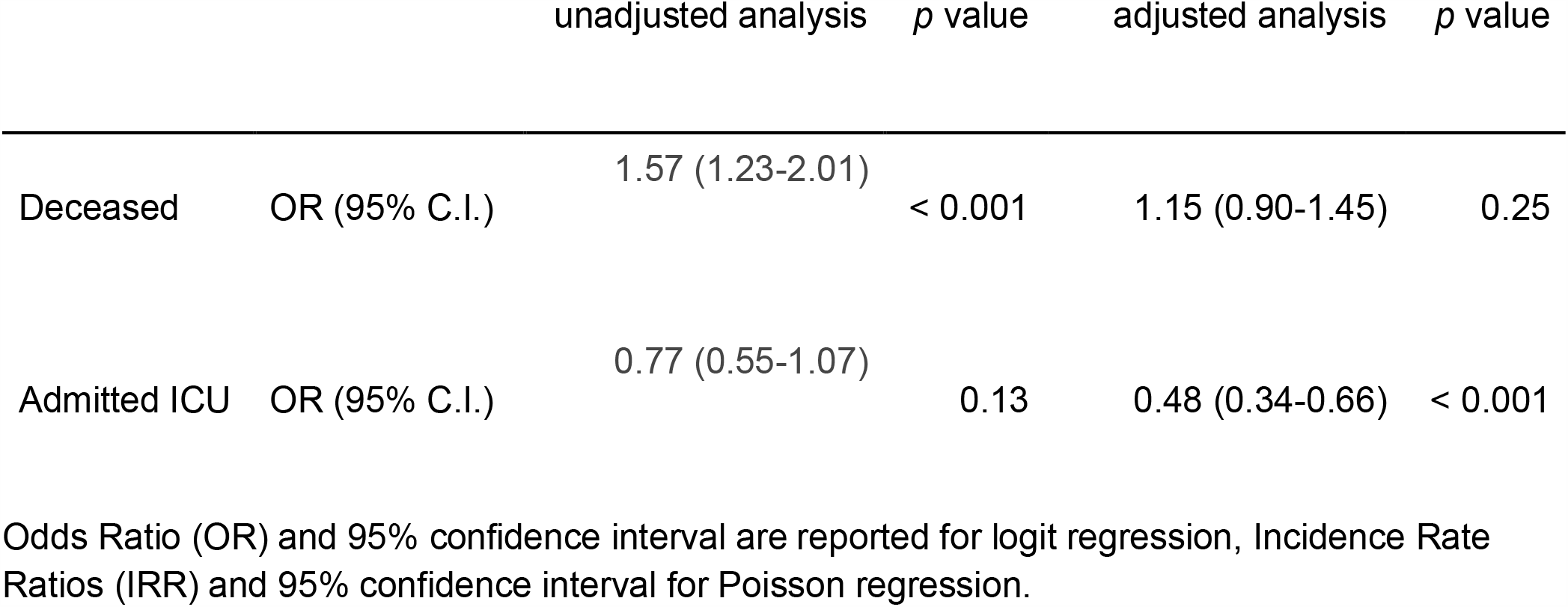

## Discussion

Treatment with corticosteroids did not affect hospital mortality in patients with COVID-19 after balancing for confounders. Since their use was first proposed as a treatment strategy for ARDS,^12,13^ corticosteroids have been at the center of controversy in the scientific community.

Some evidence points towards a beneficial effect of corticosteroids in ARDS.^12,14–16^ Recently Villar et al.^17^ showed that early therapy with dexamethasone could reduce the duration of mechanical ventilation and the overall mortality in patients with established moderate-to-severe ARDS, possibly because early administration could have an effect on the systemic immune response involved in ARDS pathophysiology. Some evidence casts doubts on the appropriateness of corticosteroid therapy: a large multicenter randomized clinical trial^18^ showed no benefit in ARDS and found an increased risk of mortality when treatment began late in the course of the disease. Moreover, corticosteroid use has been associated with complications such as increased infection rate,^19^ hyperglycemia,^20,21^ hypernatremia^22^ and ICU acquired weakness.^23^

At the beginning of the COVID-19 pandemic, caution was advised when prescribing corticosteroids, due to the possibility of a more prolonged viral shredding and reduced viral clearance.^24^ The off-label use of steroids became increasingly popular after the first study about a possible decrease of mortality in COVID-19 ARDS with the use of corticosteroids was published.^5^ Nevertheless, this study suffered some relevant limitations.

Corticosteroids therapy was assessed in a small cohort of ARDS patients (84 patients), and 50 out of them (60%) received methylprednisolone. More relevantly, data were shown without any balancing for possible confounders.

The crude mortality in our study was higher in patients treated with corticosteroids than in patients who did not receive them. The risk of in-hospital death did not differ between treated and non treated patients when adjusted for patients’ characteristics, which were similar to the ones described in previous studies.^25–27^ The lack of effect of steroids on crude mortality has been previously reported in COVID-19 patients.^28,29^ Recently a pre-print, not peer-reviewed manuscript of a randomized clinical trial (RCT) called RECOVERY trial was made available. This trial reports a 3% reduction of hospital mortality with the use of corticosteroids in COVID-19 patients. We hope that the peer reviewed results and conclusions of this study will be made available as soon as possible, so as to have a better understanding of the effects of corticosteroids in COVID-19.^30^ Nonetheless data from randomized clinical trials should always be compared to data from observational studies, because it might suffer from limited external validity. In many occasions, randomized trials on septic patients were not subsequently confirmed and their importance was reduced with time, including what concerned the use of steroids.^31–34^

A positive finding associated with the use of steroids was a reduced ICU admission rate compared to patients not receiving them. Corticosteroids can improve compliance and hypoxemia in ARDS patients,^18^ reducing the indication for ICU admission in our hospital during the COVID-19 pandemic, which was triggered by hypoxemia severity and dyspnea. In this sense, therapy with corticosteroids could be useful in reducing pressure on the Intensive Care Units in times of limited resources, as during the COVID-19 pandemic, when ICU bed occupancy had to be rationalized.

The impact of steroid therapy cannot be assessed without considering the used dose. In our patients the median equivalent dose of dexamethasone was 10 mg/die. Recent data showed that a low dose corticosteroids, defined ad lower than 1 mg·kg^-1^·day^-1^ of prednisone (equivalent to 0.15 mg·kg^-1^·day^-1^ of dexamethasone),^9^ had no effect on mortality, whereas higher doses were independently associated with and increase risk of death in patients with severe COVID-19.^35^ The weight in our patients was 77 (17) kg, which means that the average daily dose was about 0.13 mg·kg^-1^·day^-1^. Therefore the finding that low doses of steroids have no impact on mortality is confirmed by our data. The study suffers three main limitations. First, it is a retrospective analysis of prospectively collected data. Second, it is a single center study and the findings might be valid for centers with a similar setting and volume of COVID-19 admissions. Finally, propensity score with overlap method was used to balance covariate across the two treatment cohorts, so the effect of known covariate associated with mortality was taken into account and more robust conclusions could be drawn.^36^ Nevertheless the effect of unmeasured confounders or could not be excluded. In light of this limitation, meta-analyses taking into account data from this and other observational studies are needed.

In conclusion, treatment with corticosteroids was not associated with in-hospital mortality in patients admitted for COVID-19, when balancing for possible confounders. A possible advantage of corticosteroids was to reduce ICU admission, which could be useful in reducing pressure on the Intensive Care Units in times of limited resources, as during the COVID-19 pandemic.

## Data Availability

All patients deidentificated data are aviable

https://github.com/filippo1985/impact_of_AZT-HDC

## Data availability

The datasets generated during and/or analysed during the current study are available from the corresponding author on reasonable request.

## Ethics approval and consent to participate

The referral Ethics Committee (Comitato Etico di Brescia) approved the study and waived the need for informed consent from individual patients, due to the retrospective nature of the study. All research was conducted in accordance with Declaration of Helsinki.

## Consent for publication

Not applicable.

## Author contributions

All authors contributed to the data acquisition. F.A., G.N. and F.F. drafted the manuscript. F.A. designed and supervised the study. All authors contributed substantially to editing, revising and finalizing the manuscript before submission. All the authors approved the final manuscript.

## Additional Information

The study was not funded by any grants. The authors declare no competing interests.

